# Absence of SARS-CoV-2 antibodies in pre-pandemic plasma from children and adults in Vietnam

**DOI:** 10.1101/2021.07.12.21260379

**Authors:** Nguyen Van Vinh Chau, Le Nguyen Thanh Nhan, Lam Anh Nguyet, Nguyen Thi Kha Tu, Nguyen Thi Thu Hong, Dinh Nguyen Huy Man, Dinh Thi Bich Ty, Le Nguyen Truc Nhu, Lam Minh Yen, Truong Huu Khanh, Ngo Ngoc Quang Minh, Nguyen Thi Han Ny, Danielle Anderson, Lin-Fa Wang, H. Rogier van Doorn, Nguyen Thanh Hung, Tran Tan Thanh, Guy Thwaites, Le Van Tan, OUCRU COVID-19 research group

## Abstract

We tested pre-pandemic (2015-2019) plasma samples from 148 Vietnamese children, and 100 Vietnamese adults at high risk of zoonotic infections, for antibodies against SARS-CoV-2 nucleocapsid and spike proteins. None was positive, indicating no prior serological cross-reactivity with SARS-CoV-2 that might explain the low numbers of COVID-19 in Vietnam.

## Main text

Severe acute respiratory syndrome coronavirus 2 (SARS-CoV-2) emerged in late 2019, and is the cause of the ongoing coronavirus disease 2019 (COVID-19) pandemic. Yet, according to the World Health Organization, countries in the Western Pacific region, including Vietnam, have so far reported only a small fraction of the global COVID-19 cases. One hypothesis is that there may be pre-existing immunity among the population in the region through exposure to SARS-CoV-2 related viruses. Knowledge of this might help explain why the burden posed by SARS-CoV-2 and the incidence of COVID-19 cases vary significantly across the world. It may also further shed light on the natural course of the infection.

A total of 148 young Vietnamese children with hand, foot and mouth disease [1], and 100 Vietnamese adults were included for analysis of antibodies responses against SARS-CoV-2. The latter was those in close contact with domestic and/or wild animals and was thus at high risk for zoonotic infections as detailed elsewhere [2]. Of the 148 children, 8 children had RT-PCR confirmed evidence of current infection with either human coronavirus NL63 (HCoV-NL63) (n=2) or human coronavirus OC43 (HCoV-OC43) (n=6), and one adult had HCoV-OC43 detected in an earlier sample by RT-PCR. For the 100 adult cohort participants, we used plasma samples collected at baseline and two years later [2]. For children, we used 148 admission plasma from each participant, and two available convalescent plasma samples from 2/8 human coronavirus positive individuals. We included 350 plasma samples in total for the analysis. We extracted information about demographics, occupation and animal contact from the metadata of the aforementioned original studies.

We measured antibodies against two main immunogens (the nucleocapsid (N) and spike (S) proteins) of SARS-CoV-2 using two well-validated sensitive and specific serological assays, namely Elecsys Anti-SARS-CoV-2 assay (Roche, Germany) [3] and SARS-CoV-2 Surrogate Virus Neutralization Test (sVNT) (GenScript, USA) [4]. The former is an electrochemiluminescence immunoassay using recombinant N protein for qualitative detection of pan immunoglobulin (Ig) (including IgG) against SARS-CoV-2. The latter is a surrogate assay for measuring spike protein receptor binding domain (RBD)-targeting neutralizing antibodies (RBD-targeting NAbs) [4]. These two assays detected SARS-CoV-2 antibodies in plasma samples from 11/11 Vietnamese patients with PCR-confirmed SARS-CoV-2 infection collected 2 – 3 weeks after diagnosis (data not shown). The institutional review board of collaborating hospitals in Vietnam and the Oxford Tropical Research Ethic Committee approved the clinical study.

The 248 study participants came from various geographic locations in Southern Vietnam (Table 1 and Figure 1). They were all enrolled in the clinical studies between March 2013 and July 2019 (before the COVID-19 pandemic). Of the adult participants, farmers were predominant (n=42), followed by animal slaughterers (n=32) and animal health workers (n=26). The 148 children all had hand foot and mouth disease, and included 89 females and 59 males; the median age was 18 months.

**Table 1:** Demographics and animal contacts of the study participants

| Variables | Children, N=148 | Adults, N=100 |
| --- | --- | --- |
| Age<sup>#</sup> | 18 (1 - 153) | 44 (21 – 72) |
| Female/male | 89/59 (1.5) | 32/68 (0.47) |
| Occupation, n (%) |  |  |
| Farmers | NA | 42 (42%) |
| Slaughters | NA | 32 (32%) |
| Animal health workers | NA | 26 (26%) |
| HCMC origin | 62 (41.89%) | 0 |
| Other provinces | 86 (58.11%) | 100 (100%)<sup>r</sup> |
| Collection period | June, 2015 – July, 2019 | March, 2013 – September, 2016 |
| Activity/event at risk of zoonotic infection |  |  |
| Raising domestic animals | NA | 63 (63%) |
| Raising wild animals | NA | 16 (16%) |
| Bitten by animals | NA | 19 (19%) |
| Bloody injuries<sup>*</sup> | NA | 42 (42%) |

**Note to Table 1:** <sup>#</sup>in months, median (range) for children and in years, median (range) for adults
<sup>r</sup>50 from Dong Thap province and 50 from Dak Lak province
Domestic animals include dogs, chicken, pigs, cats, duck, muscovy duck, pigeons, cattle, geese, goat, rabbits, buffalo and turkeys.
Wild animals include wild pigs, deers, porcupines and monkeys.
<sup>\*</sup>Bloody injuries while working with animals.

**Figure 1:**
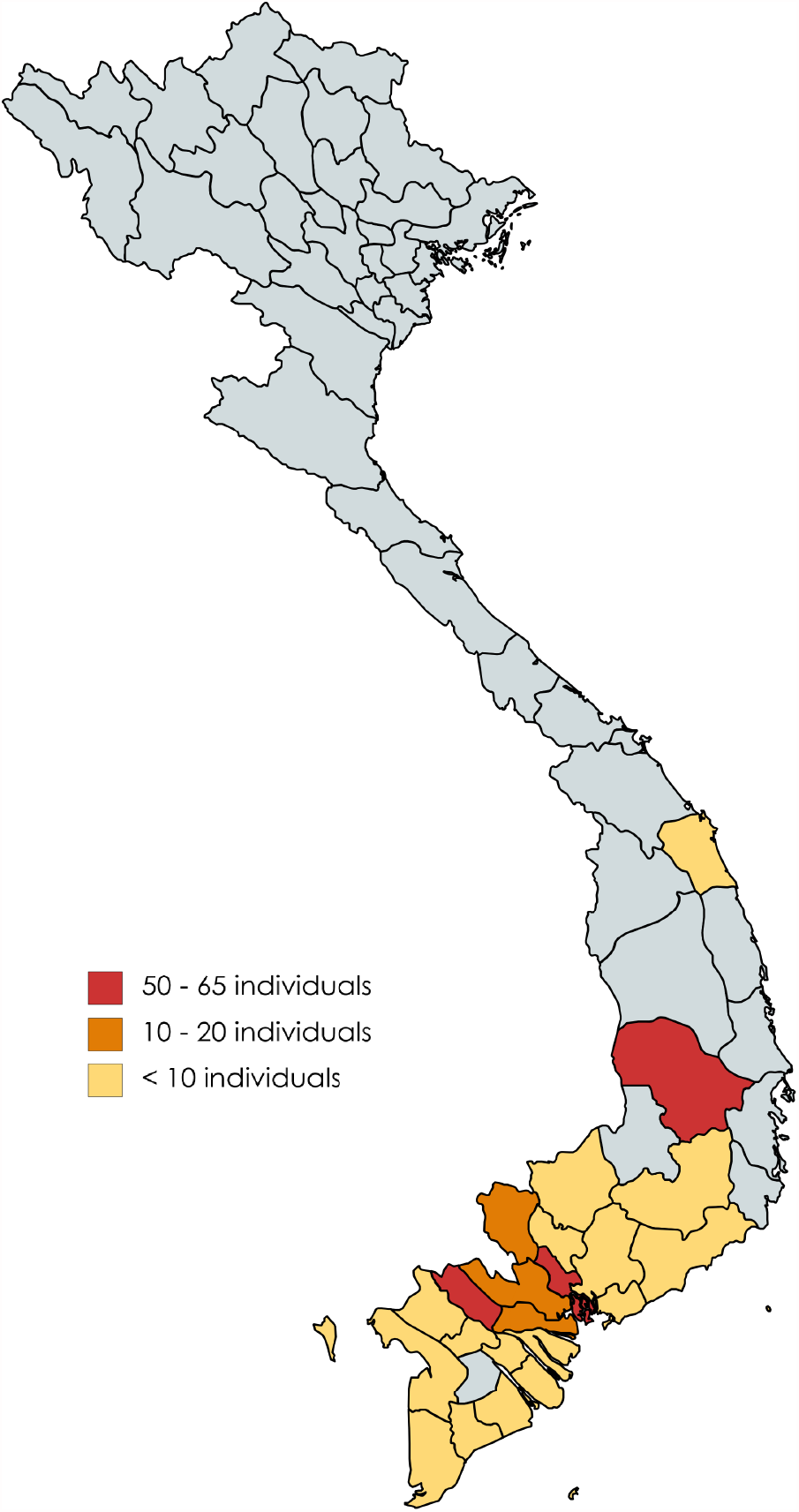
Map showing the geographic distributions of the study participants. (Inset map; https://mapchart.net).

None of the 350 plasma samples tested had detectable antibodies against N protein SARS-CoV-2. Additionally, RBD-targeting NAbs were not detected in 240 available plasma samples from the 100 adults and 38 children (including 10 from 8 positive for HCoV-NL63 or HCoV-OC43).

Using in-house immunofluorescence assays, a recent study demonstrated that antibodies against spike or nucleocapsid proteins were detected in 19% (n=105) and 14.1% (n=99) pre-pandemic plasma samples collected in Tanzania and Zambia, respectively [5]. Cross reactivity was strongly correlated with the presence of pre-existing antibodies against HCoV-NL63. Most recently, SARS-CoV-2 cross reactive antibodies, especially neutralizing antibodies, were also detected in pre-pandemic plasma from SARS-CoV-2 un-infected individuals in the UK [6].

Human coronaviruses cause the common cold worldwide with sero-prevalence increasing with age [7]. Thus, it is likely that in addition to the 9 individuals of the present study with evidence of human coronavirus infection by reverse transcription polymerase chain reaction, a proportion of the study participants were also exposed to these coronaviruses some time in the past. Therefore, previous exposure to known human coronaviruses alone might not determine the observed cross reactivity in pre-pandemic plasma. Other possible contributing factors include the difference in assay performance and/or the heterogeneities between the study populations, which merits further research.

Both cellular and humoral immunities are two major components of host responses. The former was not explored in the present study due to the unavailability of peripheral blood mononuclear cells. Of note, SARS-CoV-2-reactive T cells were detected in 20 to 50% of blood collected from unexposed individuals from various geographic locations (Germany, Singapore, the Netherlands, the United States and the United Kingdom) [8-10]. The correlation with protection of these pre-existing immunities remains unknown.

In summary, antibodies against SARS-CoV-2 nucleocapsid and spike proteins were not detected in 350 pre-pandemic Vietnamese plasma samples. Future studies should look at pre-existing B cell and T cell memory in pre-pandemic samples, which might further shed light on the pathogenesis of the infection.

## Data Availability

All data are presented in the ms.

## Acknowledgements

This study was funded by the Wellcome Trust of Great Britain (106680/B/14/Z and 204904/Z/16/Z). The serology work at Duke-NUS is supported by grants from NMRC, Singapore (STPRG-FY19-001 and COVID19RF-003).

We thank members of the VIZIONS consortium and staff from the Departments of Health and Preventative Medicine Centers (PMC) of Dak Lak and Dong Thap provinces for provision of samples, and thank VIZIONS cohort members in Dak Lak and Dong Thap provinces for their participation in the study.

We are indebted to Ms Le Kim Thanh and Ms Pham Thi Hong Anh for their logistic support. We thank the patients for their participations in this study. We would also like to thank Mr Dinh Khac Hieu for his laboratory support with the ELISA analysis.

## OUCRU COVID-19 Research Group

### Hospital for Tropical Diseases, Ho Chi Minh City, Vietnam

Nguyen Van Vinh Chau, Nguyen Thanh Dung, Le Manh Hung, Huynh Thi Loan, Nguyen Thanh Truong, Nguyen Thanh Phong, Dinh Nguyen Huy Man, Nguyen Van Hao, Duong Bich Thuy, Nghiem My Ngoc, Nguyen Phu Huong Lan, Pham Thi Ngoc Thoa, Tran Nguyen Phuong Thao, Tran Thi Lan Phuong, Le Thi Tam Uyen, Tran Thi Thanh Tam, Bui Thi Ton That, Huynh Kim Nhung, Ngo Tan Tai, Tran Nguyen Hoang Tu, Vo Trong Vuong, Dinh Thi Bich Ty, Le Thi Dung, Thai Lam Uyen, Nguyen Thi My Tien, Ho Thi Thu Thao, Nguyen Ngoc Thao, Huynh Ngoc Thien Vuong, Pham Ngoc Phuong Thao, Phan Minh Phuong

### Oxford University Clinical Research Unit, Ho Chi Minh City, Vietnam

Dong Thi Hoai Tam, Evelyne Kestelyn, Donovan Joseph, Ronald Geskus, Guy Thwaites, H. Rogier van Doorn, Ho Van Hien, Huynh Le Anh Huy, Huynh Ngan Ha, Huynh Xuan Yen, Jennifer Van Nuil, Jeremy Day, Joseph Donovan, Katrina Lawson, Lam Anh Nguyet, Lam Minh Yen, Le Nguyen Truc Nhu, Le Thanh Hoang Nhat, Le Van Tan, Sonia Lewycka Odette, Louise Thwaites, Maia Rabaa, Marc Choisy, Mary Chambers, Motiur Rahman, Ngo Thi Hoa, Nguyen Thanh Thuy Nhien, Nguyen Thi Han Ny, Nguyen Thi Kim Tuyen, Nguyen Thi Phuong Dung, Nguyen Thi Thu Hong, Nguyen Xuan Truong, Phan Nguyen Quoc Khanh, Phung Le Kim Yen, Sophie Yacoub, Thomas Kesteman, Nguyen Thuy Thuong Thuong, Tran Tan Thanh, Tran Tinh Hien, Vu Thi Ty Hang

## ABOUT THE AUTHOR

Dr Nguyen Van Vinh Chau is the Director of the Hospital for Tropical Diseases in Ho Chi Minh City, Vietnam. He is a frontline health care worker of the COVID-19 pandemic

